# Vaccination inequality and social determinants in Brazil during the COVID-19 pandemic

**DOI:** 10.1101/2025.10.01.25337110

**Authors:** Rahyja Teixeira, Gabriel S. Mouta, Jady S. M. Cordeiro, Patrícia C. S. Balieiro, Alexandre V. Silva-Neto, Daniel B. Castro, André L. C. B. Pinto, Antônio A. S. Balieiro, Antonio J.L.Costa, Vanderson S. Sampaio

## Abstract

Brazil’s vast geographic and socioeconomic diversity has contributed to heterogeneous responses to the COVID-19 pandemic, particularly regarding vaccine distribution. This ecological study examined inequality in COVID-19 vaccine administration across 5,568 Brazilian municipalities from 2021 to 2023. Vaccination coverage was estimated using the vaccination coefficient, and inequality was assessed via the Gini index. We further analyzed associations between vaccine distribution inequality and socioeconomic indicators, including the Social Vulnerability Index (SVI), and the national health infrastructure metric PQAVS-03, using beta regression models. Results indicated that northern states such as Roraima, Pará, and Amazonas consistently exhibited the highest Gini coefficients, reflecting marked disparities in vaccine allocation, while southern states like Rio Grande do Sul and Espírito Santo demonstrated more equitable distribution. SVI was significantly associated with vaccine inequality in 2021 and 2022 (p < 0.05), but this association weakened in 2023, suggesting a temporal trend toward greater equity. PQAVS-03 showed no statistically significant association in any year. These findings underscore the persistent impact of structural social determinants on vaccine access during public health emergencies, while also highlighting the potential of Brazil’s public health system to reduce disparities over time.

## Introduction

Brazil is a continental country with significant socio-economic disparities, which can lead to different patterns and outcomes of disease across its states^1^. The COVID-19 pandemic highlighted the spatial and temporal differences in terms of incidence, vaccination, and case-fatality among Brazilian States, reflecting the country’s social, economic, cultural, and structural inequalities. These disparities make it challenging to implement a one-size-fits-all solution for the entire national territory, which requires health policies adapted to regional particularities^1–3^.

Social determinants have a direct influence on the incidence and spread of infectious diseases^4^. Social conditions such as income, human capital, and urban infrastructure —often measured by indicators such as the Human Development Index (HDI) and the Social Vulnerability Index (SVI)—are directly related to COVID-19 and have a significant impact on the disease’s incidence, mortality, and case-fatality rates^5,6^.

Vaccination was the main solution to the COVID-19 pandemic^7^. The National Immunization Plan (PNI) implemented the national COVID-19 vaccination campaign to meet the needs of different population groups, taking into account the epidemiological, social, and risk-related aspects, promoting vaccination as a public health strategy^8^. In addition to providing traditional vaccines, the PNI faced unprecedented challenges, including the mass supply of newly developed vaccines, misinformation, and implementing vaccination strategies in hard-to-reach areas. It also had to confront social inequalities that influenced the population’s access to health services^9,10^.To monitor active vaccination rooms registered in the National Register of Health Establishments (CNES), the government created an important indicator for vaccine coverage surveillance - (PQAVS-3), which tracks the number of registered active vaccination rooms^11^.

Measuring health inequality in the context of vaccine distribution across different population groups in Brazil is crucial. This inequality can be quantified using ranking-based measures such as the Gini coefficient^12^, a commonly used metric to assess inequality in distribution. In the context of COVID-19 vaccination, the Gini coefficient to quantify inequalities in the distribution of COVID-19 vaccines across municipalities within each Brazilian state, aiming to capture how evenly or unevenly vaccines were allocated during the national immunization campaign. In this way, it indicates the relationship between the share of the population and the quantity of vaccines distributed. This allows for a more accurate analysis of distribution equity. The global Gini coefficients for COVID-19 vaccines were 0.91 and 0.88 in 2021^13^.

This study aimed to analyze inequalities in the application of COVID-19 vaccines across municipalities within Brazilian states from 2021 to 2023, and to assess their association with social vulnerability and local vaccination infrastructure. The study is based on the hypothesis that Brazilian states with higher social vulnerability, represented by higher SVI values, experience greater inequality in COVID-19 vaccine distribution. This hypothesis seeks to highlight the role of structural social determinants in equitable access to vaccination, contributing to our understanding of regional disparities in Brazil’s pandemic response.

## Materials and methods

An ecological study was conducted from January 2021 to December 2023. The population comprised individuals residing in Brazil, covering the 26 states and the Federal District, totaling 5,568 municipalities. Population data were obtained from the Brazilian Institute of Geography and Statistics (IBGE) (https://sidra.ibge.gov.br/home/pms/brasil). COVID-19 vaccination records were extracted from the Information System of the National Immunization Program (SI-PNI) (http://pni.datasus.gov.br/), and social indicators were obtained from the Human Development Atlas (http://atlasbrasil.org.br/2013/pt/) and made available through DataSUS.

The COVID-19 vaccination coefficient was calculated by dividing the total number of individuals vaccinated per year by the resident population per municipality for that year. (Total population was used to enable general comparisons across regions, regardless of age group or vaccine eligibility criteria.). To assess vaccine distribution inequality among States, the Gini coefficient of vaccination was calculated per Brazilian state. This was done using the cumulative population proportions and cumulative vaccinated proportions across municipalities, according to the following equation 1, where Xi represents the cumulative fraction of the total population up to the group, ordered from the smallest to the largest vaccination coefficient, and Yi represents the corresponding cumulative vaccination fraction.

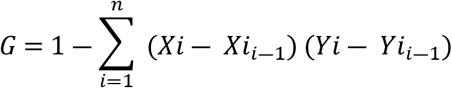

**Equation 1**‐Calculation of the vaccination Gini index

The municipality of Brasília was excluded from this analysis due to the absence of comparable groups. The Gini coefficient ranges from 0 to 1, where 0 represents perfect equality and 1 indicates maximum inequality^14^.

Two other key indices for estimating social determinants are the Social Vulnerability Index (SVI) and the Municipal Human Development Index (MHDI). The MHDI is a composite indicator that assesses the level of human development in a given location, comprising three dimensions: longevity, education, and income. It ranges from 0 to 1, with values closer to 1 indicating higher human development^15^. The SVI, in turn, estimates the degree of vulnerability and social exclusion a population faces. It also includes three dimensions: urban infrastructure, human capital, and income and employment, with values ranging from 0 to 1; the closer to 1, the higher the degree of social vulnerability^16^.

To select the social determinants, a multicollinearity analysis was conducted on the variables: SVI, SVI-Infrastructure, SVI-Human Capital, SVI-Income and Employment, MHDI, MHDI-Income, and PQAVS-03. A scatter and correlation matrix analysis revealed significant relationships among the various socioeconomic indicators. A high correlation was observed between IVS – Human Capital and SVI – Income and Employment (r = 0.922), as well as between MHDI and MHDI – Income (r = 0.976). IVS also showed high correlation with IVS – Human Capital (0.903) and SVI – Income and Employment (0.885). Negative correlations were found between MHDI and SVI – Human Capital (–0.922), MHDI and SVI (–0.816), and MHDI – Income and SVI – Human Capital (–0.947), indicating that regions with higher social vulnerability tend to register lower human development levels. Meanwhile, the PQAVS-03 indicator showed no significant correlation with most variables (maximum of 0.194), based on these findings, SVI and PQAVS-03 were selected as independent variables for the regression model (Figure 1).

**Figure 1.**
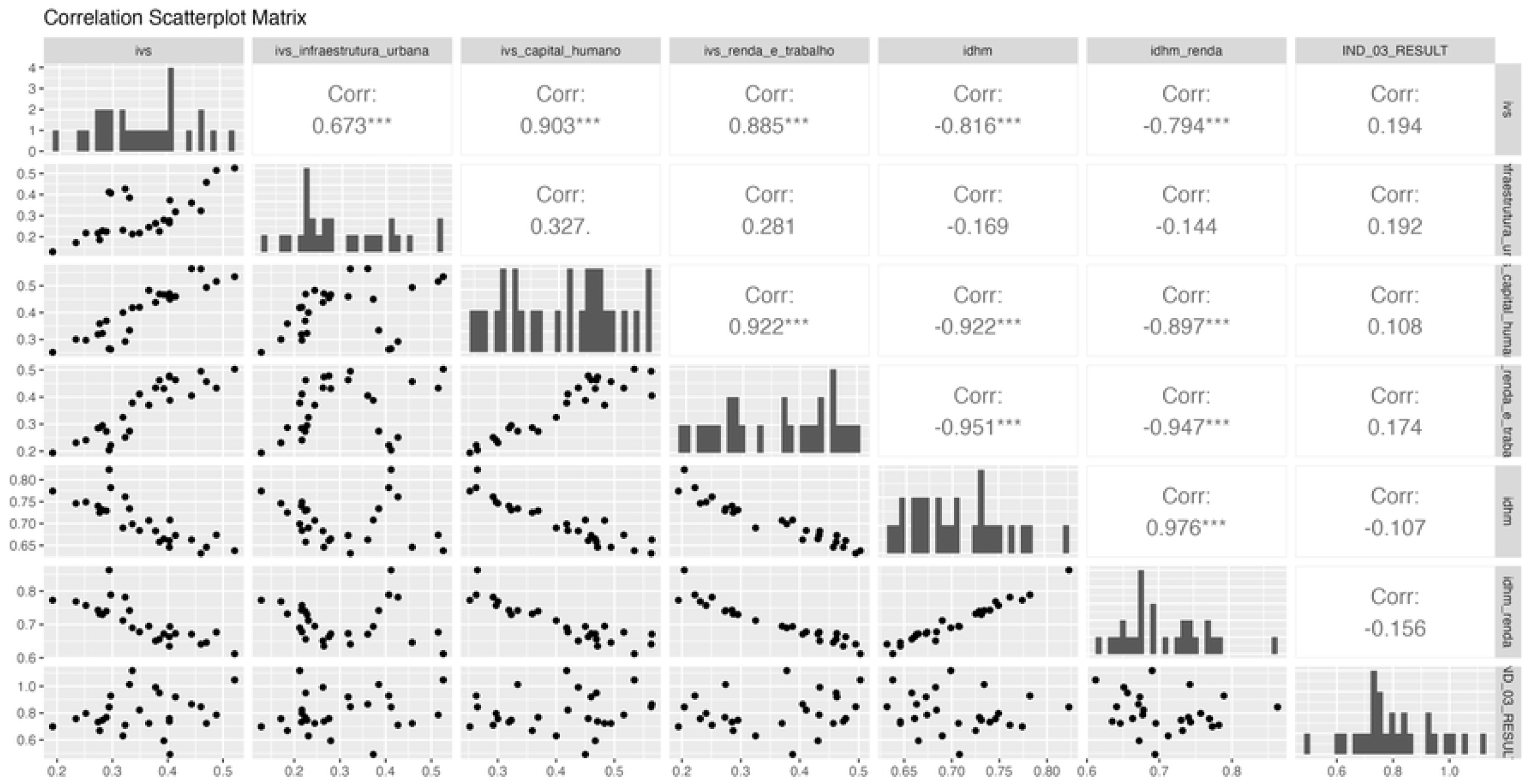
Correlation matrix of the social determinants: SVI, SVI-Infrastructure, SVI-Human Capital, SVI-Income and Work, HDIM, HDIM-Income, and PQAVS-03.

Following variable selection, a beta regression model was used to analyze the relationship between the Social Vulnerability Index, PQAVS-03, and the Gini vaccination coefficient of the states (Table S1). A significance level of 0.05 (5%) was adopted, and all confidence intervals used throughout the study were constructed at the 95% level. The beta regression model is suitable for situations where the dependent variable is continuous and constrained to the (0, 1) interval^17^.

Statistical data analysis was performed using R^18^ and RStudio^19^ software, along with Microsoft Excel Office 2010. Spatial and temporal distribution was visualized using thematic maps developed in the QGIS 3.14 environment. Cartographic data for states and municipalities were obtained from the GIS file of Brazil’s municipal grid for 2020, available on the IBGE website (https://www.ibge.gov.br/). Thematic maps georeferenced vaccination and Gini coefficients, with class divisions based on quartile intervals, using color gradients—lighter tones for lower rates and darker tones for higher ones.

This study used secondary public domain data in which subjects cannot be identified. Therefore, approval from a Research Ethics Committee was not required.

## RESULTS

The distribution of vaccination coefficients shows that the state of São Paulo stands out with the highest rates over the analyzed years, with municipalities such as Reginópolis, Fernão, Igaraçu do Tietê, and Nantes consistently appearing among the top performers. In contrast, states in northern Brazil, especially Pará and Tocantins, recorded the lowest vaccination coefficients. Municipalities such as Cumaru do Norte, Jacareacanga, Senador José Porfírio, Anapu, and São Félix do Xingu registered the lowest rates in different years. Surprisingly, the municipality of Pacaraima, in the state of Roraima, showed a notable increase in its vaccination coefficient.

The municipalities with the highest vaccination coefficients during the study were Pacaraima (7.01), Reginópolis (6.24), Fernão (5.43), Nantes (4.82), Igaraçu do Tietê (4.62), and Águas de São Pedro (3.02). Except for Pacaraima, located in the north, all the others are located in São Paulo state. The municipalities with the lowest vaccination coefficients were Cumaru do Norte (0.42), Matões do Norte (0.52), Japurá (0.53), Jacareacanga (0.54), Senador José Porfírio (0.54), and São Félix do Xingu (0.70), most of them located in northern Brazil (Figure 2).

**Figure 2.**
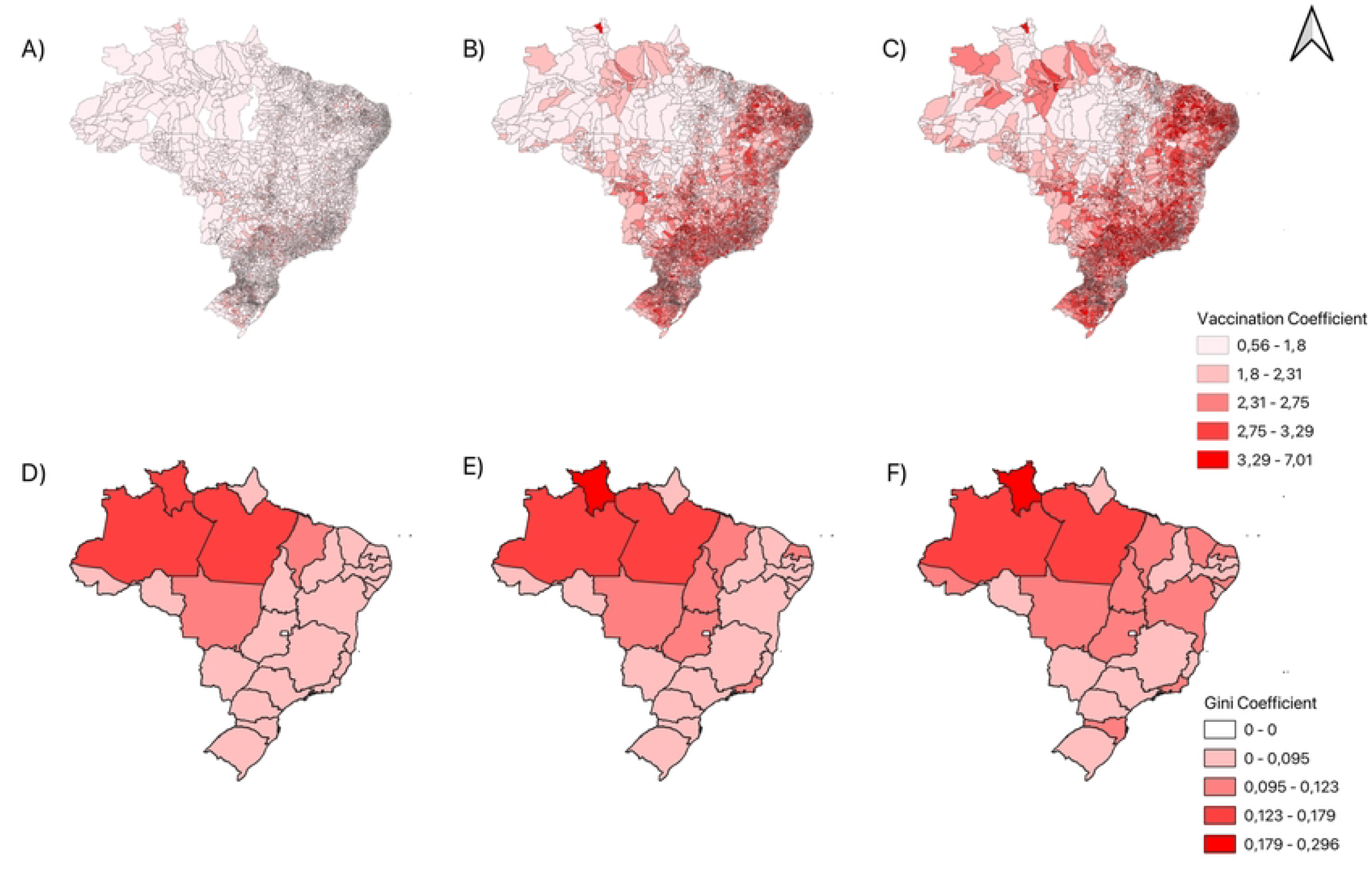
A) Vaccination coefficient of municipalities in the Brazilian territory in 2021; B) Vaccination coefficient of municipalities in the Brazilian territory in 2022; C) Vaccination coefficient of municipalities in the Brazilian territory in 2023;D) Gini coefficient of state-level vaccination in the Brazilian territory in 2021; E) Gini coefficient of state-level vaccination in the Brazilian territory in 2022; F) Gini coefficient of state-level vaccination in the Brazilian territory in 2023

Regarding the Gini coefficient of vaccine distribution, the States of Roraima, Pará, and Amazonas presented the highest levels of inequality in vaccine distribution over the years, while Rio Grande do Sul, Espírito Santo, and Sergipe presented the most balanced distribution.

The states with the highest Gini coefficients of vaccine in 2021 were Roraima (0.16), Pará (0.14), Amazonas (0.13), Maranhão (0.10), and Mato Grosso (0.09). In 2022, the highest rates were again found in Roraima (0.24), Pará (0.16), Amazonas (0.15), Mato Grosso (0.11), and Maranhão (0.11). In 2023, the same five states maintained the highest coefficients: Roraima (0.29), Pará (0.17), Amazonas (0.16), Mato Grosso (0.12), and Maranhão (0.12). These results represent the highest levels of vaccine distribution inequality.

The states with the lowest Gini coefficients in 2021 were Rio Grande do Sul (0.061), Paraíba (0.064), Sergipe (0.065), Espírito Santo (0.065), and Rondônia (0.070). In 2022, they were Rio Grande do Sul (0.066), Espírito Santo (0.069), Sergipe (0.071), and Rondônia (0.077). In 2023, the states with the lowest Gini coefficients were Rio Grande do Sul (0.074), Espírito Santo (0.075), Sergipe (0.079), Rondônia (0.080), and Amapá (0.084) (Figure 2) (Table 1).

Based on the analysis of the scatter and correlation matrix, the selection of variables for the regression model was grounded in the relationships identified among the indicators. The Social Vulnerability Index (SVI) showed significant correlations among the determinants, standing out as a central element in explaining the socioeconomic conditions analyzed. The inclusion of PQAVS-03 in the regression analysis aimed to explore potential latent relationships. The selection of SVI and PQAVS-03 for statistical modeling sought to understand how social vulnerability impacts vaccination equity.

In the univariate analysis, which examines the relationship between a dependent variable (the Gini coefficient of vaccination) and an independent variable (SVI and PQAVS), the results showed that in 2021, social vulnerability was strongly associated with vaccination inequality (p = 0.0081), while PQAVS showed no statistical relationship (p = 0.925). In 2022, the influence of SVI on vaccination inequality decreased (p = 0.054) but remained relevant. PQAVS-03 continued to have no effect (p = 0.963). In 2023, the relationship between vaccination inequality and SVI nearly disappeared (p = 0.0638), suggesting that vaccine distribution may have become more equitable over time, while PQAVS-03 remained non-significant (p = 0.984) (Table 1).

In the multivariate analysis, in 2021, SVI had a significant association with vaccination inequality (p = 0.005), while PQAVS-03 showed no influence (p = 0.479). In 2022, SVI continued to be significantly associated with inequality in vaccination (p = 0.047), and PQAVS-03 remained without effect (p = 0.641). In 2023, the influence of SVI on vaccination inequality virtually disappeared (p = 0.871), while PQAVS-03 continued to show no effect (p = 0.711). The regression analysis indicated that each one-point increase in the Social Vulnerability Index (IVS) was associated with an increase in the vaccination Gini index of approximately 1.70 in 2021, 1.58 in 2022, and 1.65 in 2023. In contrast, a one-point increase in PQAVS-03 corresponded to a decrease in the Gini index of approximately 0.25 in 2021, 0.21 in 2022, and 0.18 in 2023. These results suggest that higher social vulnerability is linked to greater inequality in vaccination coverage, while higher PQAVS-03 values are associated with reduced inequality (Table 2).

**Table 2.**
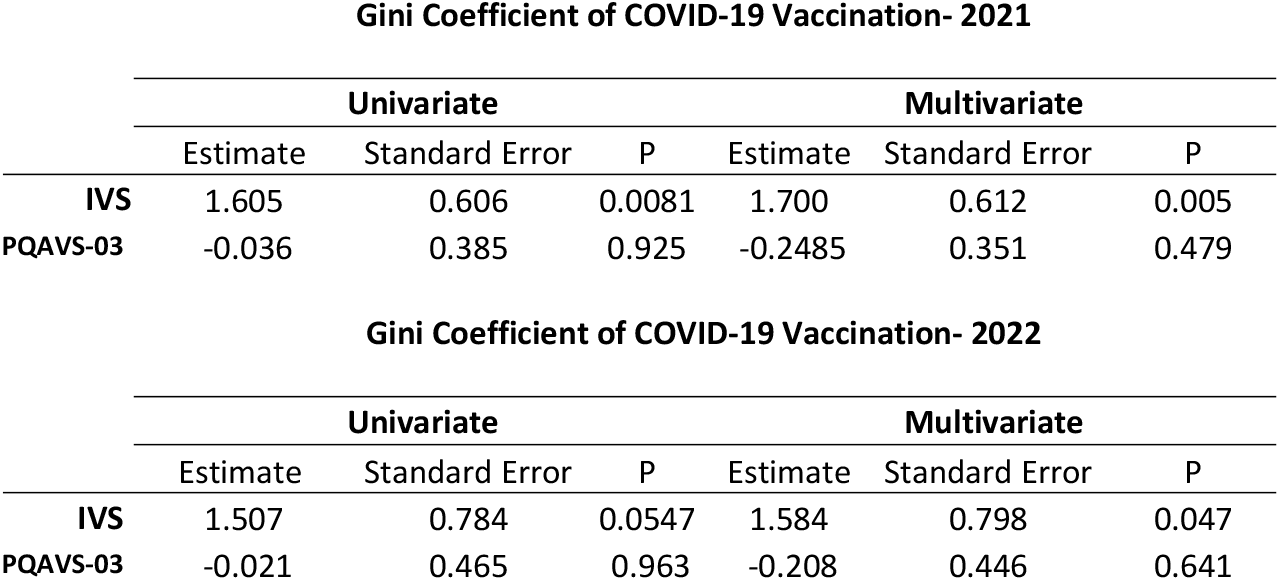
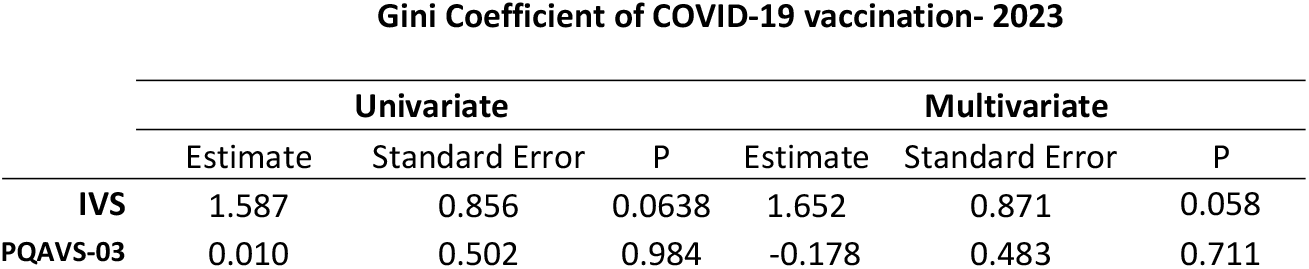
Univariate and multivariate beta regression of the COVID-19 vaccination coefficient and the Social Vulnerability Index (SVI) and Health Indicator of Vaccination Coverage (PQAVS-03).

Gini Coefficient of COVID-19 Vaccination- 2021
|  | Univariate |  |  | Multivariate |  |  |
| --- | --- | --- | --- | --- | --- | --- |
|  | Estimate | Standard Error | P | Estimate | Standard Error | P |
| **IVS** | 1.605 | 0.606 | 0.0081 | 1.700 | 0.612 | 0.005 |
| **PQAVS-03** | -0.036 | 0.385 | 0.925 | -0.2485 | 0.351 | 0.479 |

Gini Coefficient of COVID-19 Vaccination- 2022
|  | Univariate |  |  | Multivariate |  |  |
| --- | --- | --- | --- | --- | --- | --- |
|  | Estimate | Standard Error | P | Estimate | Standard Error | P |
| **IVS** | 1.507 | 0.784 | 0.0547 | 1.584 | 0.798 | 0.047 |
| **PQAVS-03** | -0.021 | 0.465 | 0.963 | -0.208 | 0.446 | 0.641 |
|  | Estimate | Standard Error | P | Estimate | Standard Error | P |
| **IVS** | 1.587 | 0.856 | 0.0638 | 1.652 | 0.871 | 0.058 |
| **PQAVS-03** | 0.010 | 0.502 | 0.984 | -0.178 | 0.483 | 0.711 |

## Discussion

The spatial distribution analysis of vaccination Gini coefficients in Brazil reveals significant inequalities among regions. The South, particularly municipalities in the state of São Paulo, stands out with the highest vaccination rates over the years, reflecting greater coverage and health-care infrastructure. By contrast, states in the North, especially Pará and Tocantins, exhibit the lowest coefficients, underscoring limitations in access and logistical challenges. Despite this scenario, there are positive exceptions, such as in Roraima, where a marked increase in vaccination rates was observed, indicating isolated advances even in historically disadvantaged areas. This outcome may be a response to intensified vaccination efforts prompted by the influx of Venezuelan migrants during that period^20,21^.

Boing and colleagues likewise described geographic, socioeconomic, and demographic disparities in COVID-19 vaccine coverage in Brazil during the first two years of the vaccination campaign, reporting significant regional inequalities in which the poorest coverage was found in the North as well as parts of the Center-West and Northeast^22^. Another study corroborates these findings, showing that the South and Southeast recorded significantly higher vaccination levels, whereas the North and Northeast recorded the lowest coverage; the same study spatially identified clusters of low vaccination strongly associated with precarious social indicators, such as low MHDI and low expected years of schooling^23^.

The present study also demonstrates that social vulnerability is associated with vaccination inequality. Regions with higher poverty rates, lower educational attainment, and poor health infrastructure tend to register significantly lower vaccination levels, especially in contexts of urban infrastructure and human capital. The North and Northeast of Brazil were the regions with the greatest inequality in vaccine distribution; these same regions present the highest Social Vulnerability Index and municipalities with low or medium MHDI, revealing historical structural inequality^24^. According to Bastos and colleagues, vaccine coverage tends to be lower in areas with a low Human Development Index, reflecting structural inequalities that directly affect public health^25^. This scenario is also observed in other countries, where determinants of vaccine distribution are influenced by socioeconomic indicators. Social inequalities shape the population’s access to health services^26^.

There is no association between the availability of vaccination rooms and vaccination inequality. A study conducted by Thakore and colleagues examined the relationship between vaccination site density, social vulnerability, and vaccination rates in the United States^27^. The results indicated that although vaccination site density was associated with vaccination rates, this relationship varied according to the level of social vulnerability in the communities^28^. Another study by Strully et al.^29^ showed that social vulnerability is linked to lower vaccination coverage - an increase of 10 percentage points in the Social Vulnerability Index was associated with a 0.87 percentage point decrease in the vaccination rate. Another study highlights that social vulnerability is strongly associated with lower COVID-19 vaccination rates in Texas, even in the presence of vaccination sites^30^. On the other hand, the study demonstrates that vaccine distribution has become progressively more equitable over time, which may be related to the strength of Brazil’s universal healthcare system (SUS) and the robustness of its National Immunization Program (PNI)^31,32^.

This study has limitations. The population estimates used to calculate the vaccination coefficient are projections based on the last available census at the time of the study, conducted in 2010. Although a new census has already been released, the most up-to-date data was not available at the time of the analysis. Similarly, the socioeconomic data is also from the 2010 census, the latest year available. The risk of inaccuracy due to the gap between the current period and the last census was minimized by grouping municipalities according to sociodemographic and health indicators in many of this study’s analyses. Furthermore, both the population and socioeconomic data are official government statistics provided by the Brazilian Ministry of Health. During the COVID-19 vaccination campaign, the SI-PNI faced limitations in recording data on administered doses. To minimize the impact of potential reporting delays on coverage calculations, we used the dataset updated in October 2024, which includes all doses administered up to December 2023.

## Conclusion

Inequality in COVID-19 vaccine distribution in Brazil was closely linked to social vulnerability, particularly in the early years of the campaign. Despite persistent regional disparities, a trend toward greater equity was observed over time, underscoring the role of the SUS and the National Immunization Program in reducing inequities. These findings highlight the need for public policies that incorporate social determinants of health to ensure equitable access in future public health emergencies.

## Data Availability

Population data were obtained from the Brazilian Institute of Geography and Statistics (IBGE) (https://sidra.ibge.gov.br/home/pms/brasil). COVID-19 vaccination records were extracted from the Information System of the National Immunization Program (SI-PNI) (http://pni.datasus.gov.br/), and social indicators were obtained from the Human Development Atlas (http://atlasbrasil.org.br/2013/pt/) and made available through DataSUS.

## Supporting information

**S1 Figure 1**. Correlation matrix of the social determinants: SVI, SVI-Infrastructure, SVI-Human Capital, SVI-Income and Work, HDIM, HDIM-Income, and PQAVS-03.

**S2 Figure 2**. A) Vaccination coefficient of municipalities in the Brazilian territory in 2021; Vaccination coefficient of municipalities in the Brazilian territory in 2022; C) Vaccination coefficient of municipalities in the Brazilian territory in 2023;D) Gini coefficient of state-level vaccination in the Brazilian territory in 2021; E) Gini coefficient of state-level vaccination in the Brazilian territory in 2022; F) Gini coefficient of state-level vaccination in the Brazilian territory in 2023.

**S3 Table 1**. Variables selected for the final regression model. The table presents Gini coefficients (2021–2023), 2010 Social Vulnerability Index (SVI), and 2021 PQAVS-03 scores for Brazilian states. Gini values reflect income inequality, with higher values indicating greater inequality. The SVI represents social vulnerability on a scale from 0 to 1, where higher values indicate greater vulnerability. PQAVS-03 scores correspond to the performance of the health surveillance action qualification program-component 3 (Programa de Qualificação das Ações de Vigilância em Saúde). Data for the Federal District (Distrito Federal) are not available for Gini coefficients.

**S4 Table 2**. Univariate and multivariate beta regression of the COVID-19 vaccination coefficient and the Social Vulnerability Index (SVI) and Health Indicator of Vaccination Coverage (PQAVS-03).

## Acknowledgement

The authors would like to acknowledge the support provided by the Data Science Laboratory at the Fundação de Medicina Tropical Dr. Heitor Vieira Dourado for their technical assistance and collaboration during the development of this study.

## References

1. Karaye, I. M. & Horney, J. A. The Impact of Social Vulnerability on COVID-19 in the U.S.: An Analysis of Spatially Varying Relationships. Am. J. Prev. Med. 59, 317–325 (2020).

2. Souza, C. D. F. de, Paiva, J. P. S. de, Leal, T. C., Silva, L. F. da & Santos, L. G. Spatiotemporal evolution of case fatality rates of COVID-19 in Brazil, 2020. J. Bras. Pneumol. 46, e20200208 (2020).

3. Rodrigues, W. et al. Social, Economic, and Regional Determinants of Mortality in Hospitalized Patients With COVID-19 in Brazil. Front. Public Health 10, (2022).

4. Bambra, C. Pandemic inequalities: emerging infectious diseases and health equity. Int. J. Equity Health 21, 6 (2022).

5. Maroko, A. R., Nash, D. & Pavilonis, B. T. COVID-19 and Inequity: a Comparative Spatial Analysis of New York City and Chicago Hot Spots. J. Urban Health Bull. N. Y. Acad. Med. 97, 461–470 (2020).

6. Coelho, F. C. et al. Assessing the spread of COVID-19 in Brazil: Mobility, morbidity and social vulnerability. PLOS ONE 15, e0238214 (2020).

7. World Health Organization. The COVID-19 vaccination response: Country experiences, best practices, and lessons.. Geneva: WHO; 2022.

8. Ministério Da Saúde / Conselho Nacional de Saúde.

9. Viana, P. V. de S., Gonçalves, M. J. F. & Basta, P. C. Ethnic and Racial Inequalities in Notified Cases of Tuberculosis in Brazil. PLOS ONE 11, e0154658 (2016).

10. Ministério da Saúde. Plano Nacional de Operalização da Vacinação contra a COVID-19. (2022).

11. Caderno de Indicadores PQA-VS 2023 Anexos I e II da Portaria no 233, de 09 de março de 2023.

12. Kawachi, I., Subramanian, S. & Almeida-Filho, N. A glossary for health inequalities. J. Epidemiol. Community Health 56, 647–652 (2002).

13. Tatar, M. et al. COVID-19 vaccine inequality: A global perspective. J. Glob. Health 12, 03072.

14. Gastwirth, J. L. The Estimation of the Lorenz Curve and Gini Index. Rev. Econ. Stat. 54, 306–316 (1972).

15. Atlas do Desenvolvimento Humano no Brasil.

16. Atlas da vulnerabilidade social nas regiões metropolitanas brasileiras. (Ipea, 2015).

17. Ferrari, S. & Cribari-Neto, F. Beta Regression for Modelling Rates and Proportions. J. Appl. Stat. 31, 799–815 (2004).

18. A language and environment for statistical computing. Vienna: R Foundation for Statistical Computing, 2024. R: The R Project for Statistical Computing. https://www.r-project.org/.

19. RStudio Team. RStudio: Integrated Development for R. https://posit.co/.

20. Moreno-Serra, R. et al. Healthcare access, quality and financial risk protection among displaced Venezuelan women living in Brazil: a cross-sectional study. Lancet Reg. Health - Am. 37, 100830 (2024).

21. Venezuelan migration in Northern Brazil: a system dynamics approach for the internalization program | Journal of Humanitarian Logistics and Supply Chain Management | Emerald Publishing. https://www.emerald.com/jhlscm/article/13/3/293/219549/Venezuelan-migration-in-Northern-Brazil-a-system.

22. Boing, A. F., Boing, A. C., Barberia, L., Borges, M. E. & Subramanian, S. V. The Brazilian vaccine divide: How some municipalities were left behind in the Covid-19 vaccine coverage. PLOS Glob. Public Health 3, e0002493 (2023).

23. Boing, A. C. et al. Spatial clusters and social inequities in COVID-19 vaccine coverage among children in Brazil. Ciênc. Saúde Coletiva 29, e03952023 (2024).

24. Desenvolvimento humano e desigualdades regionais nos municípios brasileiros. ResearchGate (2024) doi:10.25222/larr.555.

25. Bastos, L. S. L. et al. Primary healthcare protects vulnerable populations from inequity in COVID-19 vaccination: An ecological analysis of nationwide data from Brazil. Lancet Reg. Health – Am. 14, (2022).

26. de Oliveira, B. R. B. et al. Determinants of access to the SARS-CoV-2 vaccine: a preliminary approach. Int. J. Equity Health 20, 183 (2021).

27. Thakore, N., Khazanchi, R., Orav, E. J. & Ganguli, I. Association of Social Vulnerability, COVID-19 vaccine site density, and vaccination rates in the United States. Healthc. Amst. Neth. 9, 100583 (2021).

28. Tan, A. X., Hinman, J. A., Abdel Magid, H. S., Nelson, L. M. & Odden, M. C. Association between Income Inequality and County-Level COVID-19 Cases and Deaths in the US. JAMA Netw. Open (2021) doi:10.1001/jamanetworkopen.2021.8799.

29. Strully, K. W. & Yang, T.-C. County Social Vulnerability and Influenza Vaccine Rates: National and Local Estimates for Medicare Recipients. Am. J. Prev. Med. 62, e1–e9 (2022).

30. Mofleh, D. et al. Spatial Patterns of COVID-19 Vaccination Coverage by Social Vulnerability Index and Designated COVID-19 Vaccine Sites in Texas. Vaccines 10, 574 (2022).

31. Da Silva Costa, F., Santos, D. N. D., Freitas Da Silva Pinto, L., Cantuária Rodrigues, N. & Silva Monteiro, B. B. ESTRUTURAÇÃO DO PLANO DE IMUNIZAÇÃO NO COMBATE A COVID-19: (DES)ORDEM NOS ACORDOS ENTRE OS PODERES. Enferm. Em Foco 15, e–202419 (2024).

32. Portal FVS-RCP/AM. https://www.fvs.am.gov.br/pqavs.

